# A subnational family planning estimation tool (SFPET) for estimating contraceptive use and unmet need by subnational areas and demographic subgroups

**DOI:** 10.1101/2022.12.12.22283357

**Authors:** David Kong, Emily Claps, Clark Kirkman, Minerva Enriquez, Bryan Ressler, Laina D. Mercer, Sam Buxton, Mandy Izzo, Joshua L. Proctor

**Affiliations:** The Bill and Melinda Gates Foundation, Seattle, WA 98119

## Abstract

**Background:** Access to safe, effective, and voluntary family planning (FP) is a fundamental human right and is an important step toward achieving gender equality and economic autonomy. Global organizations have set ambitious goals to create universal access to FP services but assessing progress can be challenging, especially at the local geographic scale. Here, we present a quantitative visualization tool called the subnational family planning estimation tool (SFPET) that can be used by policy makers to monitor programmatic success at subnational levels and gain insight into trends within demographic subgroups.

**Methods:** The visualization tool builds on the statistical methods and analyses presented in earlier research to generate model-based estimates of FP indicators for 25 sub-Saharan countries at administrative units one and two. Here, we have extended this modeling approach to more demographic and geographic subgroups, integrated recently released survey data, and constructed a web-based dashboard to navigate survey and model-based estimates.

**Findings:** Significant heterogeneity in the levels and trends of FP indicators exist at subnational levels and across demographic subgroups. This tool provides the ability to efficiently navigate these model-based estimates across local regions within countries and across demographic subgroups. The tool also helps highlight regions and groups that may be falling behind the national trends as well as identify exemplars that exceed the national average.

**Interpretation:** SFPET^1^ is an interactive dashboard created to aid policymakers in assessing progress towards family planning goals by visualizing both model-based and direct survey estimates of family planning indicators at the subnational level. This framework provides insight into locations and population segments experiencing the greatest unmet need, enabling targeting and allocation of resources to achieve family planning goals.

## Introduction

Access to family planning is a powerful enabler of gender equality, providing women with the tools and autonomy for their health and reproductive preferences [1]. When women have access to a comprehensive set of reproductive health services, they and their families have an expanded set of opportunities that include pursuing additional educational and vocational opportunities [2]. In addition, being able to target, time, and achieve a preferred family size enables families’ ability to manage their economic resources for their children’s health and education [3]. However, progress toward universal access to family planning services has been much slower than expected [4,5] with recent global events highlighting the vulnerability women face in losing access to care [6,7].

Broad recognition of the importance of family planning has led to numerous programs—often with ambitious global goals [8]—to increase equity in access to these health services. From national agencies to global partnerships such as FP2030 (formerly FP2020, [8]), organizations and funders have invested heavily in setting and achieving quantitative targets while moving towards universal access to modern contraceptives. Recognized as a ubiquitous necessity by the United Nations, these family planning goals are also supported in the UN’s new Sustainable Development Goals (SDG) [9].

Success in achieving quantitative targets can be difficult to monitor and evaluate. Measuring progress— or the lack thereof—requires the collection of data through comprehensive monitoring programs on family planning indicators. The data most often utilized in the family planning research community comes from nationally representative cross-sectional surveys, such as the Demographic Health Surveys (DHS) [10], or to a lesser extent the more recently developed Performance, Monitoring, and Accountability (PMA) survey [11], which was designed to have more timely access to data. Statistical models and historical patterns of change are often combined with survey data to create estimation tools, such as the Family Planning Estimation Tool (FPET) [12]. Such modeling frameworks are used to fill in temporal gaps in the survey data and provide estimates of family planning indicators on annual scales as well as to forecast future trends. These tools have been incredibly valuable for understanding where progress has stagnated and for determining the root causes underlying contradictory data; unfortunately, many projected trends indicate that the goals set in SDG and FP2030 will not be achieved.

While the existing tool sets and modeling frameworks have been important to monitor family planning indicators and assess progress towards goals, they can fall short in several key areas. Although they excel in utilizing multiple types of surveys and data sources for individual countries, they often rely on national level survey estimates. Assessing programmatic success on a national level obscures subnational heterogeneities, and masks demographic sub-populations that may be significantly behind the national average. Previous research has highlighted the magnitude of this heterogeneity [13]. Understanding where variation and uncertainty occur greatly improves targeting of policy and interventions, which better serves under-represented sections of the population [13,14]. Further, models such as FPET assume an underlying logistic functional form to the model; while this enables forecasting of future trends, this assumption, without careful validation and parameterization, could cause overly optimistic short-term estimates of family planning indicators. In this modeling approach, we leverage a more data-driven approach: a model selection procedure is used to choose among a set of candidate statistical models that balances model fit and complexity.

The uptake of family planning services to empower women and families around the world is a complex, multi-faceted issue, requiring not just access to modern contraceptives, but an understanding of the local population’s behavior and needs. We believe this work is a quantitative step toward characterizing the local behavior of women in different regions and demographic subgroups. The SFPET tool can help ministries of health, policy-makers, and governmental and non-governmental funders in their decision-making around targeted interventions and monitoring activities.

## Methods

### The SFPET dashboard

We constructed the <u>subnational family planning estimation tool (SFPET)</u> to aid policymakers in assessing progress by visualizing both model-based and direct survey estimates of family planning indicators at the subnational level and by key demographic subgroups. This tool utilizes the methods described in [13] and expands the analysis by enabling the user to study a wider variety of scenarios. In brief, the modeling framework underlying the dashboard utilizes a Bayesian hierarchical modeling approach to leverage complex survey data without imposing underlying functional forms. This approach allows us to estimate and forecast levels and trends of family planning indicators at subnational scales.

Currently, the tool visualizes estimates of family planning indicators of unmet need, use of traditional contraceptive methods, and use of modern contraceptive methods at the first and second administration level for 25 sub-Saharan countries. Estimates are disaggregated across demographic subgroups, including age, parity, and urban-rural status, allowing the user to investigate subnational heterogeneities and highlight that the levels and trends can also vary significantly by demographic subgroup in the same region. This information-rich dashboard includes output and forecasts from 1990 – 2025 and provides the posterior median and 95% credible interval from the model in addition to the mean estimate and confidence interval directly from each survey. The user interface is customizable, such that the user can explore scenarios of interest as well as create custom comparisons to further delve into those scenarios. Simply select the country, year (range 1990 – 2025), administrative unit (1 or 2), desired FP indicator (unmet need, modern method, traditional method), scale and color scheme of the legend, and demographic subgroups. The demographic subgroups can be by age (15 -24 vs 25 and older), parity (0 vs 1 or more children), age and parity groups combined (i.e, 15-24 age group with 0 children), and age and urban-rural house location.

Once users select the desired criteria, the dashboard displays information-rich comparisons. A secondary map can be selected to compare across demographic groups, and information is provided for each administrative unit as well as for overall trends (with posterior median and 95% credible intervals of the model and mean estimates of the surveys with 95% confidence intervals displayed). All dashboard outputs can be saved and exported.

Currently users cannot input their own data. Model estimates are created by extracting survey data from more than 90 DHS surveys across 25 sub-Saharan countries. See [13], supplemental information, for specifics on data use and extraction; only countries with two or more surveys, associated GPS data, and access to individual level data were included. R-based scripts included in the <u>GitHub</u> repository perform the data extraction, analysis, and visualization of results. See Figure 1 for dashboard display.

**Fig 1:**
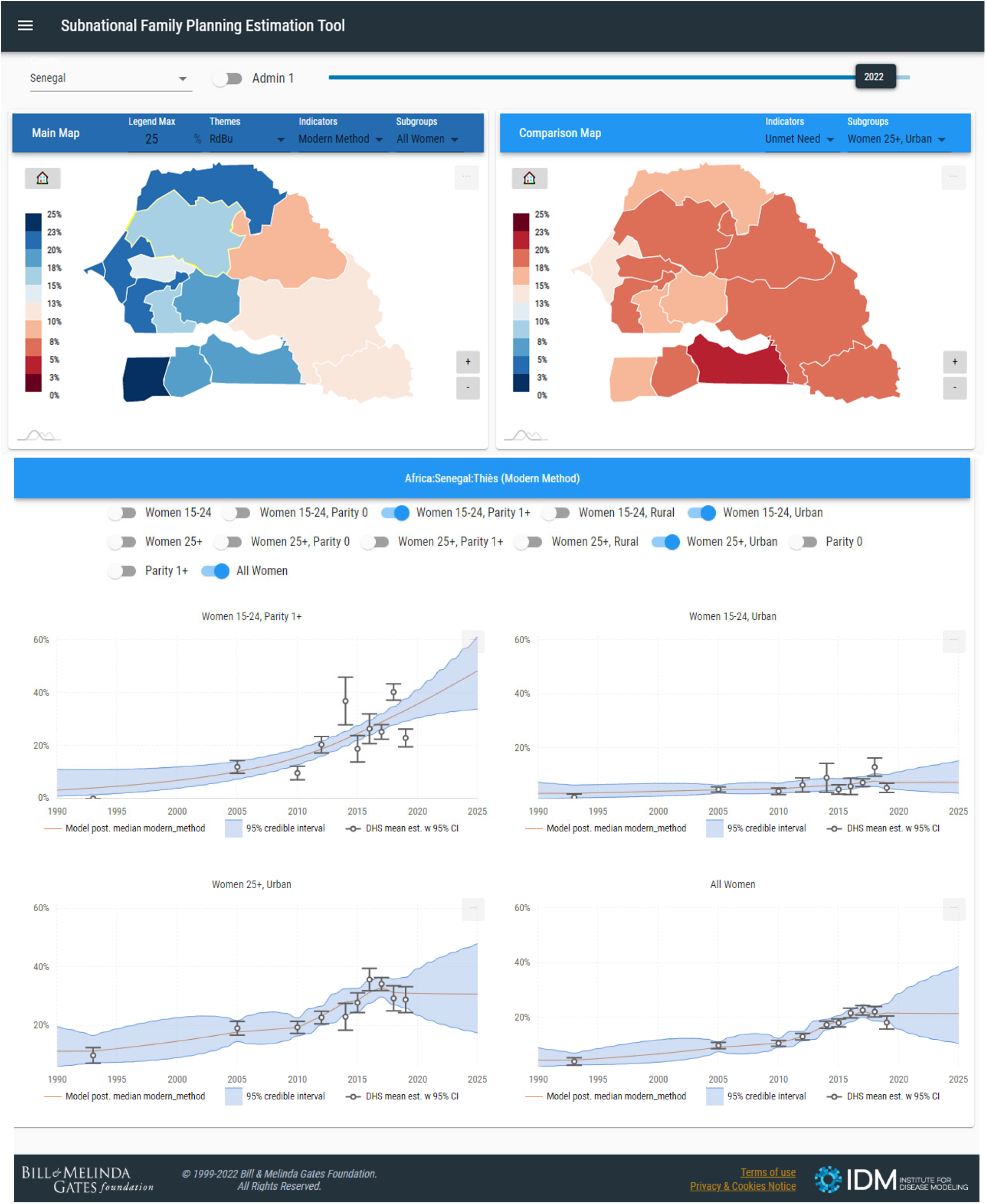
Screenshot of SFPET for Senegal with four demographic subgroups selected to assess their modern contraceptive prevalence rates from Louga.

### The models behind the dashboard

For each country and demographic subgroup, four spatio-temporal statistical models are constructed that assume an underlying rate of FP indicators with DHS survey estimates treated as measurements of these indicators with uncertainty. The direct estimates from DHS and their design-based variances were computed using the R survey package and survey design stratification variables [15]. Using a Bayesian, hierarchical modeling approach allows for the inclusion of multiple surveys, accounts for the complex survey design, and incorporates survey estimate uncertainty. Including survey estimate uncertainty is essential when estimating indicators at a subnational and demographic subgroup scale; sample sizes at this resolution can be quite small leading to noisy observations. This is a standard approach in the small area estimation statistical community [16,17]. The models include spatial and temporal independent random effects, random walk of order 1 or 2, spatially structured random effects, and temporally structured space-time interactions. Only data and trends from adjacent geographic areas are used to inform the spatial random effects. For each model fit, we use principles of model selection to balance goodness-of-fit and parsimony. The model presented in the dashboard is the most consistent across three measures of fit: log conditional predictive ordinate, Watanabe-Akaike information criterion, and the deviance information criteria; see [18] for more methodological details. It’s worth noting that these model selection criteria are useful for evaluating the fit within the time-period of available surveys for a country but the final model decision may also include assessing the realistic out-of-sample prediction trends. In these cases, we aim to iterate with stakeholders to judiciously select a model that balances realistic within and out-of-sample behavior.

### Building the dashboard

The dashboard is a visualization tool used to compare data and model output for different geographical regions. To achieve this, the platform is built by converting model data to formatted data, overlaying shape files (from the open-source GADM map and spatial data *https://gadm.org*), and building a website to display graphical information. The process is outlined in the architecture diagram below (Figure 2).

**Fig 2:**
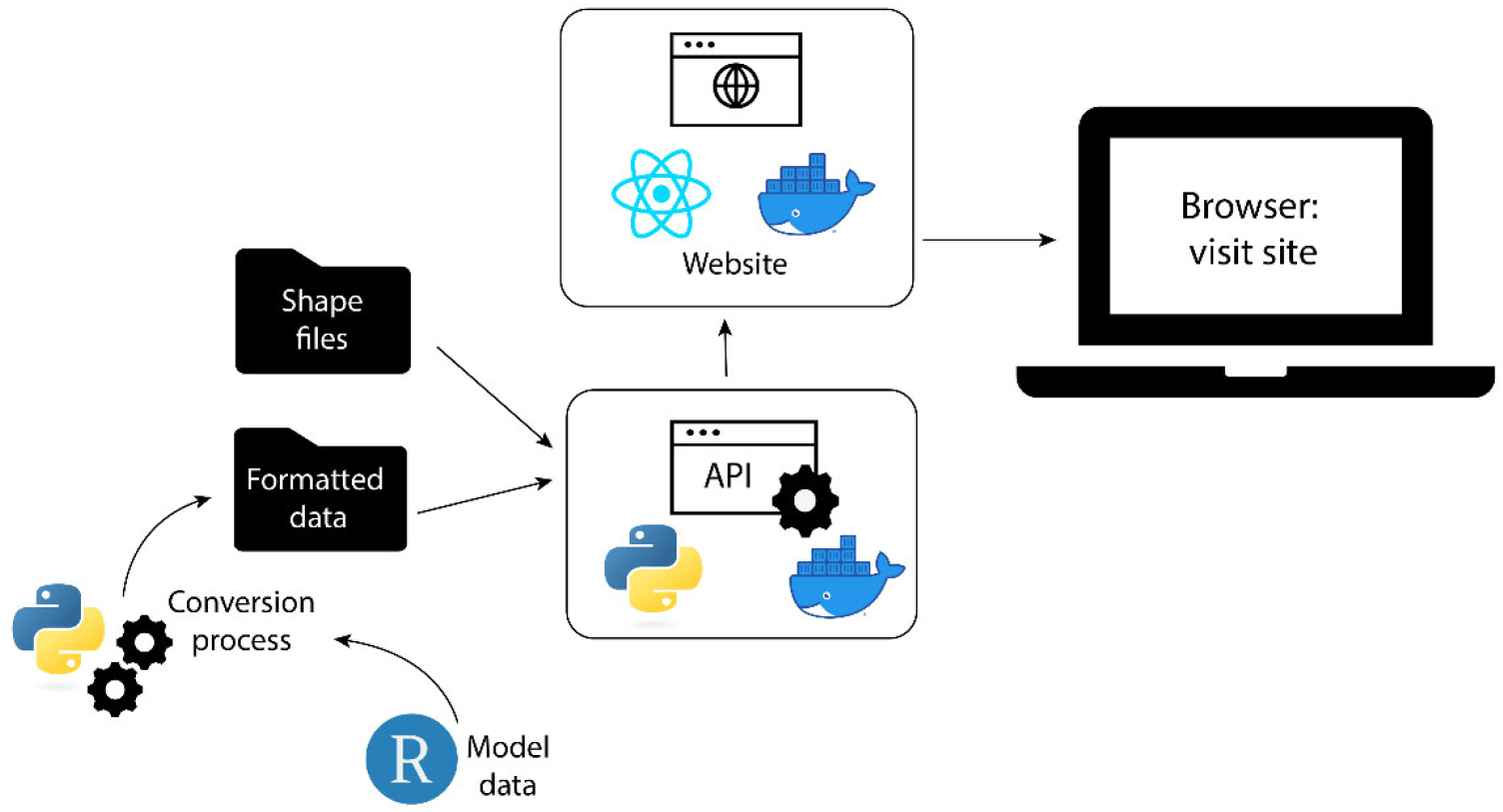
Architecture diagram demonstrating the workflow of collecting model data using R, converting it into formatted data with Python, combining it with shape files to create the API (with Python and Docker), then building the website.

The website is built from a variety of free and open-source languages with the exception of one charting library named <u>amCharts</u> which was used for plots and maps. The client website is written in Javascript, ReactJS, <u>amCharts,</u> and other open-source JavaScript libraries (see the “<u>Libraries used</u>” page on the dashboard website for complete library information). The conversion and service API layers are both written in Python. All raw model data is cleaned and packaged offline using Python scripts before it is consumed by the API. We utilize a restful API for the website to access processed model data. Both the website and service API utilize Docker containers to facilitate updating and deployment.

### Testing the dashboard

With each successive release, an automated test suite queries various facets of the API from the returned timeseries data to the label string conventions and validates the efficacy of the returned data. Once the schema of data returned by the API is validated, the tests confirm that the data is comprehensive and includes all regions for each country in the application. This is accomplished by querying the API for data from each region extracted from the shape files.

Apart from the automated testing which is run each time a change is pushed, we manually test the UI with each major release. Rather than catching missing data or bad labels, manual testing aims to catch more abstract issues with the user experience such as the focus of the map or reactiveness of the application to user input. For manual testing, we select a random sample of regions from each country, check that selecting different indicators changes the data appropriately and that the selected region and indicator is displayed in the timeseries graphs. With these two “rounds” of testing we isolate bugs at both the API and interface levels.

## Results

### Model/dashboard output

The framework that underlies this visualization tool produced estimates for family planning indicators across 25 countries in sub-Saharan Africa at the administrative 1 and 2 levels. Significant heterogeneity was found across and within the countries, both across the FP indicators of unmet need, modern contraceptive prevalence rates (mCPR), and traditional contraceptive usage rates, as well as for different demographic subgroups such as age, parity, and urban-rural groups. Detailed results can be found in the supplemental information for [14]; overall results demonstrate that while mCPR has increased at a rate of 0.75% and unmet need has decreased by 0.26% annually, these gains for all women mask the lack of progress by many subpopulations: unmet need can vary by up to 40% based on age and parity [13].

### Example scenario for the dashboard: subnational variation in unmet need for post-partum family planning (PPFP)

The SFPET visualization dashboard expands on the analysis presented in [13] and enables users to explore a variety of bespoke scenarios, delving into regions and subgroups of interest. For example, the SFPET dashboard allows users to investigate the relative rate of unmet need across women that are in different age ranges and parity (Figure 3).

**Fig 3:**
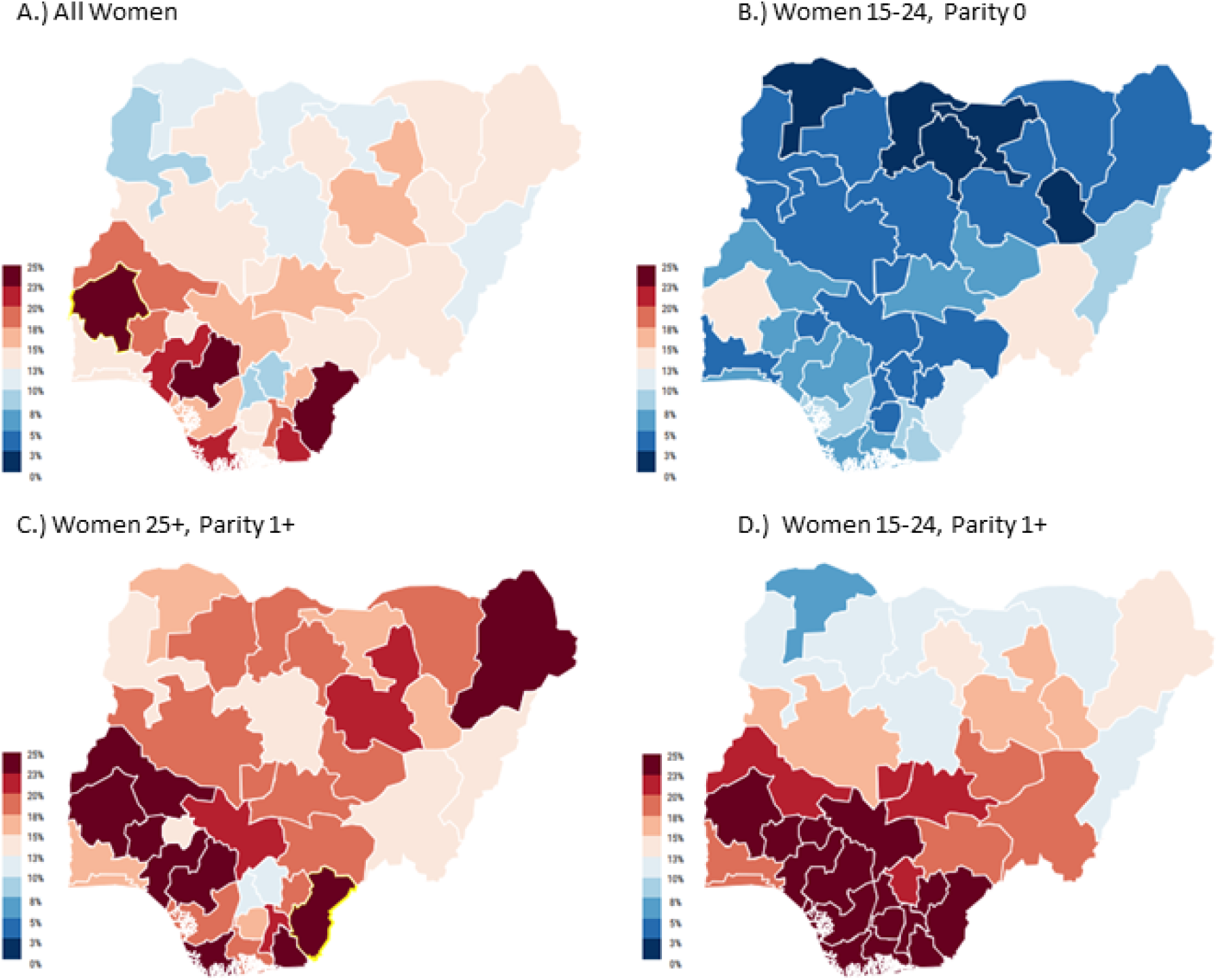
Dashboard visualization for women in Nigeria, comparing unmet need for all women (A), women aged 15-24 with no children (B), women 25 or older with children (C), and women aged 15-24 with children (D). The maps highlight the specific administrative regions with the highest rates of unmet need across these different demographic subgroups.

Examining unmet need in relation to parity is of critical importance, as post-partum family planning (PPFP) is integral for mitigating maternal and child morbidity and mortality in low-resource locations [19]. The WHO recommends spacing the birth-to-pregnancy interval to at least 2 years to prevent poor maternal, perinatal, and neonatal outcomes (such as stillbirth, premature birth or low birth weight, and neonatal and maternal mortality) [8], but it can be difficult to identify locations or age groups lacking access to PPFP. Across LMICs, birth-to-pregnancy intervals are less than 23 months long [20] with less than 61% of women using effective contraception within 24 months postpartum [21]. Parsing these groups from national data is crucial for targeted interventions.

Using the dashboard, it is easy to examine both age- and parity groups in various locations to estimate unmet need. The model-based estimates for Nigeria highlight how the rates of unmet need can be substantially different across demographic subgroups differing in age and parity. Women in many of the southern states of Nigeria that are aged 15-24 with one or more children have high rates of unmet need; in some cases, these rates are estimated to be as high as 40%. By comparison, women in the same age group with no children have much lower rates of unmet need, closer to 10%. The geographic and demographic resolution of these model-based estimates may provide data-driven insight to programmatic partners for targeting of PPFP and can help guide policymakers in determining the best location-specific intervention strategies.

## Conclusions

Universal access to safe and voluntary family planning services will be achieved by a multitude of different actions including the development of quantitative monitoring and evaluation tools such as SFPET. We have created the SFPET dashboard as an aid for researchers and policymakers to delve into nationally representative cross-sectional survey data and gain more insight into subnational trends. Disaggregating estimates of FP subnationally and by demographic group provides a framework for targeting programmatic resources and to highlight populations which may not be achieving equitable access to family planning services.

Understanding subnational variation is further enabled by exploring heterogeneities among different demographic subgroups. Parsing variables such as age, parity, and urbanicity provide valuable insight into the specific types of interventions and combinations of interventions that will be most effective. For example, examining the same age group but splitting by parity yields insight into where unmet need is the largest. Post-partum contraceptive family planning is crucial in preventing child and maternal morbidity and mortality, as mothers and children are at highest risk in the first 12 months after delivery [22]. In Nigeria’s southern states, women aged 15 – 24 with at least one child have up to 40% unmet need, almost 4x’s as high as women in the same age group with no children. These quantitative estimates by population segments can provide policymakers valuable insights as they decide on targeting programmatic interventions, such as investing in health-facility-based interventions including refresher training for providers, counseling tools, and integration with other service delivery platforms such as immunization [23,24].

The framework underlying the dashboard builds on previous tools and models to generate estimates of modern contraceptive prevalence rates, unmet need, and traditional contraceptive rates. We leverage small area estimation techniques and Bayesian hierarchical modeling to quantify uncertainty for each estimate and to incorporate sampling errors and random effects. While the results of this framework are consistent with other methodologies, this tool is the first with the ability to provide subnational estimates for 25 countries. Further, this tool is unique in its ability to enable the user to explore and visualize bespoke scenarios to examine locations and demographic subgroups of interest.

Continued progress on FP2030 and SDG ultimately requires multifaceted approaches and global commitments. Tools such as SFPET facilitate progress by identifying local heterogeneities and provide insight into what locations and demographic groups have the highest rates of unmet need. When combined with interventions such as targeted education and communication programs, public health officials can utilize quantitative frameworks such as SFPET to monitor progress and achieve quantifiable success.

## Data Availability

The data used to generate the model outputs and produce the maps in the dashboard are all publicly available. The dashboard website also has an option for directly downloading model estimates directly.

https://dhsprogram.com/

https://gadm.org

## Acknowledgements

The authors would like to acknowledge Fred Lu, Qinghua Long, Meikang Wu, and Guillaume Chabot-Couture for their support of this research over the last few years.

## Footnotes

1 https://sfpet.bmgf.io/

